# Making Invisible Infrastructure Visible: Data Landscapes Across a National Pain and Opioid Research Network through a Qualitative Assessment

**DOI:** 10.64898/2026.11.24.26368823

**Authors:** Meredith C. B. Adams, R. Andrew Taylor, Jason A. Hoppe, The HD2A Network

**Author notes:** Co-Senior authors contributed equally to this work (R. Andrew Taylor, Jason A. Hoppe). **Corresponding Author**: Meredith C. B. Adams, MD, MS, Department of Anesthesiology, Artificial Intelligence, Translational Neuroscience, and Public Health Sciences, Wake Forest University School of Medicine, Medical Center Boulevard, Winston-Salem, NC 27157; Fax: N/A.

## Abstract

**Background:** Multi-site research networks have become central infrastructure for NIH-supported health research, yet network-level coordination has focused on harmonization, common data models and FAIR principles, while rarely characterizing the heterogeneous data landscapes within a network first. The Helping to End Addiction Long-term^®^ Initiative (HEAL Initiative) Data2Action (HD2A) program comprises ten autonomously designed innovation and acceleration projects in opioid use disorder treatment and pain management, with no pre-existing network-level view of what data each project used or how it was governed.

**Objectives:** To make the HD2A data-infrastructure landscape visible by systematically characterizing the data environments of all ten projects and identifying cross-project themes, coordination opportunities, and lessons for multi-site networks.

**Methods:** Data Infrastructure Support Center staff extracted each project’s data-environment characteristics from project documentation into a structured 15-element assessment, which project investigators reviewed and corrected. Two analysts independently coded the ten validated assessments (two-pass inductive content analysis; 19-code codebook), developed six cross-project themes from the coded passages by consensus, and judged each project against them. An investigator reflection survey, summarized descriptively, contextualized the document-based findings.

**Results:** Six themes characterized the landscape: all projects integrate multi-source, cross-sector data; dashboards are the common data-to-action venue; adoption of standards-based interoperability is uneven; governance complexity increases nonlinearly with institutional partners; implementation and improvement science frameworks serve as data-quality scaffolding; and sustainability plans depend on institutional transfer. Five synergy clusters were identified. Investigators consistently described data access and governance, not analytic capability, as the binding constraint, and distinguished sustaining technical products from sustaining their use.

**Conclusions:** Making a network’s data infrastructure visible is not a substitute for harmonization but a prerequisite for deciding how and what to harmonize. HD2A’s autonomously designed projects share a common structure of constraints, creating opportunities for coordination without uniformity. The structured elicitation approach is replicable in other innovation networks.

## INTRODUCTION

Over the past two decades, multi-site research networks have become a central infrastructure for NIH-supported health research, including programs such as PCORnet, the CTSA consortium, the NIH Helping to End Addiction Long-term® Initiative (HEAL Initiative), and disease-specific clinical trial networks.^1–3^ These programs bring together investigator teams spanning institutional, geographic, and disciplinary boundaries, each working with data environments shaped by local infrastructure, institutional partnerships, and regulatory contexts.^4, 5^ The practical promise of these networks, cross-site learning, shared resources, and coordinated evidence generation, depends on their ability to make sense of the diverse data environments they encompass.^6–8^ However, the dominant model for multi-site data coordination has focused primarily on harmonization: adopting common data models, building centralized platforms, and applying FAIR principles to make data findable, accessible, interoperable, and reusable.^9, 10^

Harmonization approaches have generated substantial value, but they presuppose a step that is rarely undertaken: systematically characterizing the heterogeneous data landscapes within a network before determining how and what to coordinate.^11, 12^ When a network includes projects drawing on electronic health records, prescription drug monitoring programs, criminal justice databases, community surveillance systems, patient-reported outcomes, and qualitative stakeholder data, each source operates under distinct governance regimes, refresh cycles, and quality standards. In this context, the question of what data exist across the network, how they are governed, and where projects converge or diverge in their data infrastructure is not answered by a common data model alone.^13, 14^ Instead, it requires a different kind of work: understanding and making the network’s data infrastructure visible to the network itself.

The concept of “invisible infrastructure” has a well-established lineage in science and technology studies. Bowker and Star introduced “infrastructural inversion” as a method for foregrounding infrastructure that, by design and habit, fades into the background when it is functioning well.^15, 16^ In research networks, data environments are invisible in precisely this sense: each project’s data infrastructure works well enough for that project’s purposes, but the network lacks a mechanism for viewing the collective landscape, including where projects face similar challenges, where they have made divergent choices, and where shared investment could reduce duplicated effort.^17, 18^ This invisibility is not a failure but a structural feature of networks designed to support locally appropriate data innovation. However, it limits what the network can learn about and from itself.

The NIH HEAL Initiative’s Data2Action (HD2A) program illustrates this challenge. Funded by the National Institute on Drug Abuse beginning in 2022, HD2A comprises ten innovation and acceleration projects using real-world data to improve opioid use disorder treatment and pain management across clinical, public health, and community settings, supported by three resource centers focused on implementation science, data infrastructure, and health economics. The programmatic structure and support center architecture are described elsewhere, and a qualitative formative evaluation has been published.^19^ HD2A projects were designed with considerable autonomy in selecting their data sources, analytic approaches, and implementation frameworks, a design reflecting the program’s emphasis on locally relevant, stakeholder-driven data innovation. This autonomy meant that no mechanism existed at the network level to characterize what data each project was working with, how those data were governed, or what patterns of convergence or challenge were operating across the portfolio.

This paper reports what became visible when the Data Infrastructure Support Center (DISC), one of three HD2A resource centers, led the network in systematically examining its own data landscape. Using a three-phase qualitative study, structured data extraction from project documentation, two-pass qualitative content analysis producing a formal codebook, and an investigator reflection survey capturing experiential knowledge, we characterize the data infrastructure landscape of HD2A and identify cross-project themes, coordination opportunities, and lessons for multi-site networks. The contribution is not a new harmonization framework or common data model; instead, it provides empirical evidence about what a national health innovation network looks like when its data infrastructure is made legible, and a replicable approach for other networks seeking to move from unexamined heterogeneity to actionable intelligence about their own data environments.

## METHODS

### Setting and Study Design

We conducted a three-phase, multi-method qualitative descriptive study to characterize the data-infrastructure landscape of the HD2A network. Phase 1 was a structured extraction of data-environment characteristics from each project’s documentation using a structured assessment instrument, validated by each project’s investigators. Phase 2 was a two-pass qualitative content analysis of the ten validated assessments, yielding a 19-code codebook, six cross-project themes developed from the coded passages, and a theme-by-project mapping. Phase 3 was an investigator reflection survey, fielded after extraction was complete, that captured experiential knowledge the documentation did not surface and was used to contextualize, not to generate or revise, the themes. The study is reported per the Standards for Reporting Qualitative Research (SRQR).^20^ We took a pragmatic stance and used a qualitative descriptive design because the study’s purpose was to summarize each project’s data environment in terms close to the source documentation and in a form the network could act on, not to develop theory or interpret lived experience.^21, 22^ We chose conventional (inductive) content analysis for the same reason: prior work offers little guidance on characterizing data infrastructure across a multi-project research network, so codes were developed through close reading of the project assessments and consensus discussion between analysts, not imposed in advance.^23^

### Ethics Statement

The Wake Forest University Health Sciences Institutional Review Board reviewed this study and determined it to be exempt (Protocol #IRB00146435). No patients took part, and no patient-level data were collected or analyzed. Phases 1 and 2 used project documentation and the structured assessments derived from it. For Phase 3, investigators and staff of the ten HD2A projects received an invitation that described the survey and explained that participation was voluntary; completing the REDCap survey was taken as consent to participate. Because the study was determined to be exempt, signed consent forms were not required and were not collected. The survey asked about project-level data infrastructure and did not request personal or sensitive information. Responses were stored in REDCap on secure institutional servers accessible only to the study team and are reported in aggregate, without attribution to individual respondents.

### Research Team and Reflexivity

The study was designed and carried out by the DISC, whose funded role is to support the data infrastructure of the ten projects examined here. Both analysts (MCBA, [PI, pain/addiction informatics]; RWH, [Co-I, pain/addiction informatics]) are DISC members with established working relationships with the project teams; they were embedded in the network, not independent of it. This position gave the analysts access to project documentation and a contextual understanding of each project’s data environment. It also carried two risks: that the analysts’ support role and familiarity with the projects would shape what was extracted and how it was interpreted, and that teams might describe their data environments favorably to a center that supports them. We addressed these risks in several ways. Each project’s principal investigator and data team reviewed and corrected their own assessment before analysis, so the analyzed assessments reflect project-verified content and not DISC’s reading alone. Both analysts coded all ten assessments independently before comparing results. Every code and theme judgment was anchored in assessment text, discrepancies were resolved by consensus with a third DISC team member (CC) available to adjudicate, and decisions were logged in an audit trail. Findings are framed as coordination opportunities, not as evaluations of individual projects. Two authors (RAT, JAH) are investigators on projects included in the analysis. They did not extract data, code, or derive themes; like all project investigators, they reviewed only their own project’s assessment for accuracy, and they contributed to interpretation of the findings and revision of the manuscript.

### Qualitative Content Analysis

#### Phase 1: Structured Data Extraction

The DISC developed a structured instrument, the Data2Action Network Project Assessment Framework, comprising 15 free-text elements in five sections: a data characterization matrix (data types and sources; data volume and complexity; data quality indicators; data sharing mechanisms); an environmental success factors assessment (regulatory and legislative environment; institutional infrastructure; collaborative ecosystem factors); a project trajectory analysis (embarkation success indicators; completion success factors); an analytical checklist (pre-conditions assessment; process enablers evaluation; outcome facilitators review); and a comparative analysis framework (data type clustering analysis; environmental condition correlation; success pattern recognition). A closing field recorded an overall assessment of the project. For all ten innovation and acceleration projects, two-member DISC teams extracted data-environment characteristics from project documentation (grant applications, data-management plans, and related materials) into this instrument between November 2025 and February 2026. Extracted content was recorded as discrete free-text entries, typically a phrase or a single sentence, under each element of the instrument. Each project’s principal investigator and data team then reviewed and corrected the extracted summary to confirm accuracy. The PI-validated version of each assessment was treated as final and formed the data analyzed in Phase 2. The instrument was refined iteratively through engagement with the Research Adoption Support Center, which recommended alignment with the Consolidated Framework for Implementation Research (CFIR),^24, 25^ and the Modeling and Economic Resource Center, which advised on impact measurement.

#### Phase 2: Two-Pass Qualitative Content Analysis

The data for Phase 2 comprised the ten PI-validated structured project assessments, one per project. The grant applications, data-management plans, and related materials from which the assessments were extracted were not themselves coded; project documentation also supplied descriptive context for the Results (for example, the size of a health system or the agencies with which a project coordinates), but this material was not coded. The unit of coding was the assessment entry: each phrase or sentence the extraction team recorded in response to an instrument item. Entries were double coded when they met the inclusion criteria for more than one code, with all code assignments agreed by consensus.

The assessments were analyzed using a two-pass, inductive (conventional) content analysis.^23^ The two DISC analysts independently coded all ten assessments. In the first pass, preliminary codes were derived inductively from the entries, each assigned a brief name, a definition grounded in the assessment text, and one to three exemplar entries quoted verbatim from the assessments; no codes were imposed a priori. In the second pass, the preliminary codes were refined into a formal codebook with explicit definitions, evaluating each code for redundancy, breadth, and operational clarity. Overlapping codes were merged (e.g., NLP and interoperability standards into a single technical data-processing code), overly broad codes were split (e.g., quality measurement into external standards compliance and internal performance benchmarking), and vague definitions were operationalized with inclusion and exclusion criteria. The final codebook comprises 19 codes organized in four clusters: Infrastructure and Technical Capacity, Governance and Partnership, Measurement and Quality, and Stakeholder Engagement and Capacity, and is provided in S1 Appendix.

The two analysts then independently applied the final codebook to every entry in all ten assessments and resolved all discrepancies by discussion to consensus, with a third DISC team member (CC) adjudicating unresolved cases. Rigor was supported by independent dual coding, anchoring of every code in verbatim text, explicit inclusion/exclusion criteria, and a decision audit trail. Consensus code assignments were tabulated in a code-by-project matrix recording which codes were applied to each project and the entries that supported them.

##### Derivation of cross-project themes

After coding was complete, the two analysts read the coded passages across all ten projects together and, by consensus, developed six themes, each an interpretive statement about a pattern shared across projects. A theme could draw on passages from more than one code, and codes whose passages did not converge into a cross-project pattern relevant to the study objective remained in the codebook but were not reported as themes. For each theme the analysts wrote an operational definition stating what a project’s assessment had to contain for the theme to be considered present. Table 1 gives each theme’s definition and contributing codes.

**Table 1.** Derivation of the six cross-project themes from the 19-code codebook. For each theme, the operational definition the two analysts applied when judging a project’s assessment as present or absent, and the codebook codes whose coded entries were reviewed for that judgment.

| **Theme** | **Operational definition: the theme was marked present when the project's assessment contained at least one coded entry that…** | **Contributing codebook codes (S1 Appendix)** |
|---|---|---|
| 1. All ten projects integrate multi-source, cross-sector data | described linking or integrating data from two or more distinct source systems (e.g., electronic health record, prescription drug monitoring program, emergency medical services, vital statistics, claims, criminal-legal, or social-service data). | 1, 17, 14 |
| 2. Dashboards are the common data-to-action venue for translating data to diverse audiences | identified a dashboard or comparable interactive visual display as a primary means of delivering data to decision-makers. | 2 |
| 3. Adoption of standards-based interoperability is uneven | named a national data, terminology, or interoperability standard or a standards-based architecture (e.g., HL7, FHIR, SMART on FHIR, CDS Hooks, RxNorm, LOINC, SNOMED, standards-based application programming interfaces or computable phenotypes). Entries describing natural language processing or project-specific linkage without a named standard did not qualify. | 5, 6 |
| 4. Governance complexity increases nonlinearly with the number and diversity of institutional partners | described a formal data-governance arrangement, either a signed agreement (e.g., data sharing agreement, memorandum of understanding, data use agreement, business associate agreement) or a standing oversight or advisory structure. Differences in complexity across projects were then characterized narratively from the number and type of partner organizations named. | 3, 4 |
| 5. Implementation and improvement science frameworks serve as data-quality scaffolding | named a formal implementation science, quality improvement, or systems framework or method used to organize the project's data collection, measurement, or feedback. | 8, 9, 6 |
| 6. Sustainability plans depend on transferring data products to institutional partners | described a plan or mechanism for embedding data products or operations in, or transferring them to, an institutional or governmental partner beyond the grant period. | 13, 12 |

The two analysts then jointly judged each project as present or absent on each theme by reading that project’s entries coded to the contributing codes against the theme definition. A theme was marked present when at least one coded entry met the definition, an analyst judgment anchored in coded text rather than an automatic roll-up of code occurrence. For example, Code 5 covers both natural language processing and interoperability standards, but Theme 3 was marked present only for projects whose entries named a national standard or a standards-based architecture, so a project whose only Code 5 entries concerned natural language processing was marked absent. In the resulting theme-by-project mapping (Table 2), a blank cell indicates that the validated assessment contained no entry meeting the definition; it does not establish that the project lacks the corresponding feature.

**Table 2.** Cross-Project Theme Mapping. Presence of each theme in the ten PI-validated project assessments, judged jointly by two analysts against the theme definitions in Table 1 (N = 10 projects). ✓ = at least one coded entry in the project’s assessment met the theme definition; blank = no entry met the definition.

| Theme | Aalsma/Ray/Reda | Banta-Green | Cruden | Venkatesh | Heck-mann | Hoppe | Karnik | Marshall-Wright | Neigh-bors | Sun |
|---|---|---|---|---|---|---|---|---|---|---|
| 1. All ten projects integrate multi-source, cross-sector data | ✓ | ✓ | ✓ | ✓ | ✓ | ✓ | ✓ | ✓ | ✓ | ✓ |
| 2. Dashboards are the common data-to-action venue for translating data to diverse audiences | ✓ | ✓ |  | ✓ | ✓ | ✓ | ✓ | ✓ | ✓ |  |
| 3. Adoption of standards-based interoperability is uneven |  |  |  | ✓ |  | ✓ | ✓ |  |  | ✓ |
| 4. Governance complexity increases nonlinearly with the number and diversity of institutional partners | ✓ | ✓ | ✓ | ✓ | ✓ | ✓ | ✓ | ✓ | ✓ | ✓ |
| 5. Implementation and improvement science frameworks serve as data-quality scaffolding | ✓ | ✓ | ✓ | ✓ | ✓ | ✓ |  | ✓ | ✓ |  |
| 6. Sustainability plans depend on transferring data products to institutional partners | ✓ | ✓ | ✓ | ✓ | ✓ | ✓ |  | ✓ | ✓ |  |

#### Phase 3: Investigator Reflection Survey

After extraction was complete, we surveyed all project teams to capture tacit knowledge about data work that project documentation does not record; all ten projects participated, each submitting one response. The survey was fielded in April 2026 via REDCap and comprised ^26^ primarily open-ended items spanning the project’s most significant data challenge; governance, sharing, and access constraints; data-quality validation; sustainability (distinguishing technical sustainment from continued use); and advice for incoming projects. The summarized results and the survey items were then discussed with project investigators at an in-person network meeting in April 2026, which served as member checking of the summary. The full instrument is provided in S3 Appendix.

Survey responses were analyzed separately from the project assessments. They were not coded with the 19-code codebook and were not used to develop or revise the codebook, themes, or theme-by-project mapping. Instead, one DISC team member (CC) summarized the responses item by item, grouping similar responses and counting the number of projects that raised each issue, with the project as the unit of analysis (n = 10). The survey served to contextualize and triangulate the document-based findings: to examine whether patterns identified in the assessments were consistent with investigators’ experience, and to surface experiential knowledge that project documentation did not record.

## RESULTS

Analysis of the ten project assessments identified six themes describing how projects acquire, integrate, govern, and sustain their data environments (Table 1). Themes were not equally present across projects (Table 2). The following sections synthesize each theme, using exemplar projects to illustrate convergence and variation within the portfolio.

### Theme 1: All Ten Projects Integrate Multi-Source, Cross-Sector Data

All ten projects (identified hereafter by principal investigator surnames) integrate data from multiple, fundamentally distinct data sources, making cross linkage the defining feature of the HD2A data landscape (Codebook Codes 1, 17). This is more than simple data aggregation; projects perform complex record linkage across systems with different governance regimes, refresh cycles, and quality standards. Aalsma/Ray/Reda link criminal legal records, emergency department (ED) data, emergency medical services (EMS) interactions, and prescription dispensation across Indiana’s population, refreshing weekly and monthly. Hoppe integrates prescription drug monitoring program (PDMP), electronic health record (EHR), and mortality data across a 12-hospital system with over 3,000 providers. Sun links a 50-institution surgical registry with EHR and Medicare claims. At the community level, Banta-Green and Hood integrate EMS medical incident reports with a county-level data hub spanning jail, substance use treatment, housing, and Medicaid utilization.

Projects diverge in how they manage this integration. Several adopt standards-based identifiers and architectures (Hoppe, Karnik), while others rely on project-specific matching algorithms or manual validation. This variation likely reflects differences in institutional data maturity and the nature of data sources; clinical systems are more likely to support interoperability standards than community surveillance or criminal justice systems.

### Theme 2: Dashboards Are the Common Data-to-Action Venue for Translating Data to Diverse Audiences

Eight of ten projects identified dashboards as a primary mechanism for translating data into actionable outputs (Codebook Code 2). Aalsma/Ray/Reda’s Overdose Touchpoints Dashboard provides real-time population health data to fatality review teams. Banta-Green and Hood use EMS and mortality dashboards with heat maps for stakeholders. Venkatesh is developing an ED opioid use disorder quality dashboard within the American College of Emergency Physicians (ACEP) registry. Neighbors provides performance feedback dashboards stratified by race, ethnicity, and gender. Marshall/Wright’s CADENCE project centers on an interactive maternal-infant outcomes dashboard.^26^ Cruden was an informative exception: because existing dashboards in its setting made new dashboards problematic, the project is developing data products other than dashboards.

Dashboard audiences varied: dashboards serve county-level overdose fatality review (OFR) teams of local public health, legal, and treatment providers and people with lived experience (Aalsma/Ray/Reda), clinical quality improvement teams (Venkatesh), organizational leadership (Neighbors), and community stakeholders (Heckmann). This diversity of audiences, combined with the consistency of the dashboard form, points to dashboards as a dominant translational technology in applied health data innovation.

### Theme 3: Adoption of Standards-Based Interoperability Is Uneven

Four projects reported explicit adoption of national data standards (Codebook Codes 5, 6), though the degree of standards-based infrastructure varied substantially. Hoppe implements SMART on FHIR (Fast Healthcare Interoperability Resources) and clinical decision support (CDS) Hooks.^27^ Karnik uses Health Level ^28^ (HL7) and FHIR for unifying screening data. Venkatesh utilizes ACEP data system ontologies (RxNorm, LOINC, SNOMED) and application programming interfaces. Sun develops computable phenotypes using standard ontologies. The remaining projects describe data integration approaches relying on project-specific linkage methods, manual validation, or institution-specific dictionaries without reference to national standards.

This divergence emerged most clearly through cross-project comparison and likely reflects the projects’ starting positions relative to existing infrastructure rather than differences in analytic sophistication. Projects embedded in health systems inherit standards-based architectures; those working within EHR environments had readier access to FHIR-compatible systems, whereas community-based or multi-agency projects working with administrative data from criminal justice or public health surveillance systems must construct interoperability from heterogeneous sources.

### Theme 4: Governance Complexity Increases Nonlinearly with the Number and Diversity of Institutional Partners

All ten projects had formal governance arrangements (Codebook Codes 3, 4). Projects within a single health system (Hoppe at UCHealth; Karnik at UIC) described governance through institutional IRB processes and internal data use agreements. For example, CDS approval may require navigating multiple separate governance and quality improvement projects. Multi-institutional projects reported substantially more complex landscapes: Aalsma/Ray/Reda coordinate with the Indiana departments of Health, Corrections, State Police, and Homeland Security. Cruden maintains standard operating procedures and data sharing agreements across multiple Oregon counties and state agencies. Venkatesh sustains memoranda of understanding across ACEP, five ED sites, and a quality improvement network.

Partnership count did not merely add governance burden; it introduced qualitatively different challenges. Single-institution projects negotiate within familiar organizational hierarchies, while multi-agency projects must navigate across institutional cultures, legal frameworks, and data-sharing norms that may conflict.

### Theme 5: Implementation and Improvement Science Frameworks Serve as Data-Quality Scaffolding

Eight projects adopted formal implementation science (IS), improvement science, or systems frameworks not merely for their clinical interventions but as organizing structures for their data work (Codebook Code 8). Six used IS theories, models, or frameworks. Aalsma/Ray/Reda and Cruden use the Exploration, Preparation, Implementation, Sustainment (EPIS) process and determinants framework. Hoppe uses the Practical, Robust Implementation and Sustainability Model (PRISM) with Reach, Effectiveness, Adoption, Implementation, Maintenance (RE-AIM), a determinants and evaluation framework. Neighbors employs the Health Equity Implementation Framework. Venkatesh uses the Technology, People, Organizations, and Macroenvironmental factors framework and RE-AIM and aligns measures with National Quality Forum and Centers for Medicare & Medicaid Services guidelines. Marshall/Wright used CFIR and RE-AIM alongside interactive process mapping.^28^ The remaining two drew on improvement science (Banta-Green and Hood, continuous process improvement) or systems science (Heckmann, system dynamics modeling).^29–32^

The prevalence of these frameworks across project data environments suggests that IS and improvement science have become a de facto organizing logic for applied health data work, not only guiding clinical intervention delivery but structuring how teams think about data quality, measurement, and reporting. Projects are not treating data infrastructure and IS as separate domains; they are using these constructs to define what counts as quality data, what outcomes to measure, and how to organize feedback loops between data and practice. This integration has implications for how networks conceptualize data support: data infrastructure consultation may need to engage with these frameworks as foundational to how project teams organize their data work, rather than treating data as a purely technical concern.

### Theme 6: Sustainability Plans Depend on Transferring Data Products to Institutional Partners

Eight projects described sustainability strategies centered on transferring data products to institutional or governmental partners rather than maintaining independent research systems (Codebook Codes 13, 12). Cruden plans to transfer its data repository to the Oregon Health Authority. Aalsma/Ray/Reda leverage the Indiana Department of Health’s existing OFR support infrastructure. Banta-Green and Hood build on the PI’s role as co-executive sponsor of the Washington State Opioid Response Workgroup. Hoppe aligns CDS tools with UCHealth’s strategic plan. Venkatesh integrates measures into ACEP’s registry, which health systems use to meet Merit-based Incentive Payment System criteria. Neighbors collaborates with the Office of Addiction Services and Supports (OASAS), which provides in-kind support. Heckmann reports that community partners have acquired their own funding.

### Cross-Project Data Synergy Clusters

Cross-project analysis identified five clusters where shared infrastructure approaches could generate network-level value beyond what individual projects achieve independently. Each represents an opportunity for network-level coordination.

#### Real-Time Overdose Surveillance

Aalsma/Ray/Reda, Banta-Green/Hood, and Heckmann each developed near-real-time overdose surveillance dashboards from EMS, mortality, and ED data. Shared dashboard design standards, common data elements for overdose surveillance, and shared data refresh guidance could reduce duplication.

#### Clinical Decision Support and Standards-Based Infrastructure

Hoppe, Karnik, and Sun operate within health system environments and have adopted FHIR, HL7, CDS Hooks, or computable phenotypes. Shared validation approaches or phenotype libraries could benefit all three and produce reusable network resources.

#### Quality Measurement and Value Alignment

***Venkatesh*** and Neighbors focus on quality measurement and performance feedback in different settings (EDs and opioid treatment programs). A shared approach to quality measure specification or health equity reporting could create a transferable model.

#### Multi-Sector Administrative Data Linkage

Aalsma/Ray/Reda, Banta-Green/Hood, Cruden, and Neighbors perform cross-sector linkage spanning health, criminal justice, housing, and social services. Shared governance templates, linkage validation methods, or privacy-preserving approaches could address common bottlenecks.

#### PDMP Integration

Hoppe, Karnik, and Aalsma/Ray/Reda integrate PDMP data, facing shared challenges around interstate variability, data currency, and clinical record linkage. Shared guidance could benefit these projects and inform future initiatives.

### Investigator Reflection Survey

All ten projects responded to the reflection survey, with one response per project. Findings are reported in aggregate with the project as the unit (n = 10) and without attribution to individual projects.

#### Data challenges and governance

^26^ of ten projects named cross-system integration as their most significant data challenge, and five named governance, sharing agreements, or access permissions explicitly; the two were typically described together rather than in isolation. Investigators described several downstream consequences of constrained access: narrowed analytic scope when key linkages proved infeasible, dashboards displayed without key demographic breakdowns, county-level suppression that obscured local signal, and lengthened iteration cycles.

#### Data-quality validation

Projects with raw-data access described concrete validation procedures, including exploratory data analysis with outlier detection, inter-annotator agreement for annotated data, and Sankey diagrams of cross-system linkage. Projects without raw-data access stated that their validation capacity was bounded by what governance permits.

#### Sustainability

Several projects reported high confidence that their dashboards, pipelines, and data processes could survive the end of the award provided institutional or governmental partners absorb them, and described sustaining mechanisms already in place, including state government backing, multi-million-dollar institutional resourcing, and alignment with state-level opioid response governance. One project drew the distinction between sustaining technical products and sustaining their use explicitly: it had no concern about sustaining the technical artifacts themselves, only about whether community stakeholders would continue to use them. Projects named user training, dashboard benchmarking, and continued engagement of community partners as the activities most at risk of erosion after the grant period, and at least two reported being expected to sustain their products without the resources to do so.

#### Advice for incoming projects

When asked what advice they would offer an incoming project, five of ten projects independently named some version of planning for governance friction before planning for analysis.

## DISCUSSION

### What the HD2A Data Landscape Reveals

By systematically characterizing the data environments of all ten HD2A innovation and acceleration projects and surfacing the cross-project themes that organize them (Table 1), we produced a portrait of a network whose projects share deep structural similarities in their data challenges despite having been designed with deliberate autonomy. Five synergy clusters showed where coordination could create value across projects without requiring uniformity. A complementary investigator reflection survey, conducted with HD2A teams after the structured assessment was complete, sharpened the interpretation of several of these themes and added findings that the assessment instrument alone did not surface. Considered together, the document analysis and the reflection survey reveal not only what a national health innovation network looks like when its data infrastructure is made legible, but where the network’s most consequential constraints actually lie. Consistent with accounts of infrastructure as becoming visible chiefly at points of breakdown,^15, 16^ these constraints surfaced where project data work met governance and access friction.

### The Binding Constraint Is Governance, Not Analytics

The most consistent message from investigators, expressed across multiple reflection-survey items, is that data access and governance, not analytic capability, is the network’s binding constraint. Integration and governance challenges were typically described together, validation capacity was bounded by what governance permits for projects without raw-data access, and five of ten projects advised incoming teams to plan for governance friction before analysis. This pattern mirrors reports from cross-sectoral linkage efforts in which legal, governance, and access barriers, rather than technical capacity, limit routine data linkage,^13, 14^ and it exposes a gap in data-quality frameworks that presume analysts can inspect the underlying records.^11, 12^

This finding reframes how the network’s heterogeneity should be understood. The variation in interoperability adoption and in governance complexity is not principally a difference in technical sophistication; it reflects the context of the data systems from which projects begin. Health-system projects working within EHR environments inherit FHIR-compatible infrastructure and within-institution governance pathways; community-based and multi-agency projects working with administrative data from criminal justice, public health surveillance, EMS, or Medicaid systems must construct linkage architectures while simultaneously negotiating sharing agreements across organizations with conflicting legal frameworks. The downstream consequences investigators described are governance artifacts more than analytic ones. A key finding was that no amount of additional analytic capacity solves a governance bottleneck, and network-level investment in linkage methods alone will under-deliver if it is not paired with investment in the legal, regulatory, and relational scaffolding that determines what can be linked at all.

### Sustainability Is Not One Problem but Two

A second finding that emerged with greater clarity in the reflection survey than in the document review is that sustainability in HD2A is split between technical sustainment and use sustainment, and the human side is the weaker of the two. Several sites reported high confidence that their dashboards, pipelines, and data processes can survive the end of the award provided institutional or governmental partners absorb them, yet the concern investigators raised was whether community stakeholders would continue to use them. This distinction is consequential. The network-wide convergence on “institutional transfer” sustainability strategies presumes that the receiving institution has the technical capacity, staffing, and organizational incentive to maintain the product after handoff. The reflection survey suggests that an additional, underexamined condition must also hold, that the intended users continue to find the product worth using as practice and policy evolve.

Framed this way, sustainability planning requires two distinct strategies. Technical sustainment depends on integration with institutional or governmental data infrastructure, leadership-level resource allocation, and operational alignment with a clinical or public health workflow, mechanisms that several sites described as concretely in place, including state government backing, multi-million-dollar institutional resourcing, and alignment with state-level opioid response governance. Use sustainment depends on whether the audiences for whom the data product was built (clinicians, public health officials, community stakeholders, EMS responders) remain embedded in feedback loops that keep the product useful over time. Sites named user training, dashboard benchmarking, and continued engagement of community partners as the activities most at risk of erosion after the grant period. The implication for network design is that sustainability support should be conceptualized along both axes: technical maintenance templates and institutional partnership agreements address the first, but the second requires durable investment in stakeholder engagement, training continuity, and the measurement of actual use.

### Implications for Innovation Networks

Several implications for network design emerge from these findings. First, network-level data characterization should be initiated at the outset of a network’s operations, not retrospectively. Early characterization allows the network to identify coordination opportunities while projects are still developing their infrastructure, rather than after approaches have solidified.

Second, the convergence on dashboards across HD2A suggests that network data infrastructure should be conceptualized as an interpretation system as well as a pipeline, with investment in how data reaches the people who act on it. Dashboard design, user testing, and audience segmentation may be as important to network data infrastructure as data standards or governance. The convergence may also reflect its timing: many projects were proposed when COVID-19 dashboards were ubiquitous, and funder and reviewer expectations may have favored the form, much as current expectations favor integrating artificial intelligence. Networks should consider how such top-down expectations shape the infrastructure projects adopt.

Third, the governance complexity theme suggests that networks should differentiate their governance support based on the number and type of institutional partners a project engages, rather than providing uniform guidance. A project linking data across four state agencies faces qualitatively different governance challenges than one working within a single health system, and network support should reflect that difference.

Fourth, sustainability support should address both technical and use sustainment as distinct problems; institutional transfer agreements solve the first but not the second.

Fifth, funder mechanisms should accommodate the adaptive character of two-stage transitions such as NIH R61 to R33 grants, allowing scope and approach to evolve in response to implementation learning through instruments such as protocol amendments, administrative supplements, exploratory pre-implementation aims, and implementation-science-focused opportunities. The rapid evolution of artificial intelligence since funding began has the potential to advance projects’ analytic and technical capabilities, even as the underlying governance constraints remain unchanged.

## LIMITATIONS

### Nature of the data

*The analysis did not include* independent observation, primary qualitative data collection (e.g., interviews), or audit of data systems. Because Phase 2 coded the assessments and not the underlying source documents, the coded text reflects the extractors’ condensation of those documents, and the exemplar quotations in S1 Appendix are assessment entries, not investigators’ own words. This limits the depth of evidence for some themes and means that the characterization reflects how teams describe their data environments, which may differ from operational reality. Because all ten projects were included, the sample was a census of the network rather than one sampled to thematic saturation.

### Analyst position

The analysis was conducted by a resource center embedded in the network it studied. Investigator validation of each assessment, independent dual coding, and consensus procedures reduce, but do not eliminate, the possibility that the analysts’ support role shaped extraction and interpretation.

### Network-specific context

This study was conducted within a single network focused on opioid use disorder and pain management. Applicability to other networks, disease areas, or data contexts requires further evaluation.

### Method as infrastructure

The structured characterization approach used in this study is itself a form of data infrastructure, with its own design choices and limitations. The assessment’s section structure inevitably shaped what became visible; themes that fall outside those sections may have been underrepresented. In particular, the assessment evolved to incorporate IS constructs during the data collection period, and elements added later (such as the CFIR-aligned inner/outer setting distinction) may not have been captured consistently across all projects.

## CONCLUSIONS

Making a network’s data infrastructure visible is not a substitute for harmonization, but a prerequisite for deciding what to harmonize. Although HD2A’s data landscape was intentionally designed to be heterogeneous, its projects nonetheless share a common structure of constraints, governance as the binding limitation, dashboards as the primary translational interface, and institutional transfer as the dominant sustainability pathway. This shared structure creates concrete opportunities for coordination without requiring uniformity. The methods used here are replicable in other innovation networks. The findings suggest that what such networks most need from their support infrastructure is not better tools alone, but better visibility into the often-invisible conditions under which their tools must work.

## Acknowledgements

We would like to acknowledge Janet Kuramoto-Crawford, NIH Scientific Officer for the Data Infrastructure Support Center (DISC), for her guidance and support. We would like to acknowledge the contributions of Niranjan S. Karnik and Neeraj Chhabra under grant number R61DA057629/R33DA057629 for their contribution of contribution of participation in this project. We would like to acknowledge the contributions of Matthew Aalsma under grant number R33DA057660 for their contribution of participation in this project. We would like to acknowledge the contributions of Douglas A Colquhoun under grant number R33DA059168 for their contribution of participation in this project. We would like to acknowledge the contributions of Gracelyn Cruden under grant number R33DA059163 for their contribution of contribution of participation in this project.

## Data Availability Statement

The qualitative codebook (S1 Appendix), the survey instrument (S3 Appendix), the theme definitions with their contributing codes (Table 1), and the theme-by-project mapping (Table 2) are within the paper and its Supporting Information files. The ten structured project assessments cannot be shared publicly because they were derived from unpublished grant applications and internal project documentation that belong to the individual HD2A project teams and their institutions, not to the authors. Requests for a project’s assessment or its source documentation should be directed to that project’s principal investigator (list of projects, principal investigators and contact information: (Supplemental Files), or to Janet Kuramoto-Crawford, NIH Scientific Officer for the Data Infrastructure Support Center (DISC). Individual survey responses are not publicly available because the protocol reviewed by the Wake Forest University Health Sciences Institutional Review Board limits reporting to aggregate results and, with ten identifiable projects, individual responses could be attributed to respondents.

## Supporting information

**S1 Appendix.** HD2A qualitative codebook. The 19 codes with their cluster, definition, inclusion and exclusion criteria, and exemplar assessment entries.

**S2 Appendix.** HD2A Network Corporate Authorship

**S3 Appendix.** Blank Extraction Instrument.

**S4 Appendix.** Investigator reflection survey instrument.

**S1 Checklist.** Standards for Reporting Qualitative Research (SRQR) checklist.

## References

1. Collins FS, Koroshetz WJ, Volkow ND. Helping to End Addiction Over the Long-term: The Research Plan for the NIH HEAL Initiative. JAMA. 2018;320(2):129–30. doi: 10.1001/jama.2018.8826. PubMed PMID: 29896636; PMCID: PMC6688196.

2. Fleurence RL, Curtis LH, Califf RM, Platt R, Selby JV, Brown JS. Launching PCORnet, a national patient-centered clinical research network. J Am Med Inform Assoc. 2014;21(4):578–82. Epub 20140512. doi: 10.1136/amiajnl-2014-002747. PubMed PMID: 24821743; PMCID: PMC4078292.

3. Visweswaran S, Becich MJ, D’Itri VS, Sendro ER, MacFadden D, Anderson NR, Allen KA, Ranganathan D, Murphy SN, Morrato EH, Pincus HA, Toto R, Firestein GS, Nadler LM, Reis SE. Accrual to Clinical Trials (ACT): A Clinical and Translational Science Award Consortium Network. JAMIA Open. 2018;1(2):147–52. Epub 20180821. doi: 10.1093/jamiaopen/ooy033. PubMed PMID: 30474072; PMCID: PMC6241502.

4. Doria-Rose VP, Greenlee RT, Buist DSM, Miglioretti DL, Corley DA, Brown JS, Clancy HA, Tuzzio L, Moy LM, Hornbrook MC, Brown ML, Ritzwoller DP, Kushi LH, Greene SM. Collaborating on Data, Science, and Infrastructure: The 20-Year Journey of the Cancer Research Network. EGEMS (Wash DC). 2019;7(1):7. Epub 20190329. doi: 10.5334/egems.273. PubMed PMID: 30972356; PMCID: PMC6450242.

5. Forrest CB, McTigue KM, Hernandez AF, Cohen LW, Cruz H, Haynes K, Kaushal R, Kho AN, Marsolo KA, Nair VP, Platt R, Puro JE, Rothman RL, Shenkman EA, Waitman LR, Williams NA, Carton TW. PCORnet(R) 2020: current state, accomplishments, and future directions. J Clin Epidemiol. 2021;129:60–7. Epub 20200928. doi: 10.1016/j.jclinepi.2020.09.036. PubMed PMID: 33002635; PMCID: PMC7521354.

6. Embi PJ, Richesson R, Tenenbaum J, Kannry J, Friedman C, Sarkar IN, Smith J, members of APIPC. Reimagining the research-practice relationship: policy recommendations for informatics-enabled evidence-generation across the US health system. JAMIA Open. 2019;2(1):2–9. Epub 20190116. doi: 10.1093/jamiaopen/ooy056. PubMed PMID: 31984339; PMCID: PMC6951885.

7. McGlynn EA, Lieu TA, Durham ML, Bauck A, Laws R, Go AS, Chen J, Feigelson HS, Corley DA, Young DR, Nelson AF, Davidson AJ, Morales LS, Kahn MG. Developing a data infrastructure for a learning health system: the PORTAL network. J Am Med Inform Assoc. 2014;21(4):596–601. Epub 20140512. doi: 10.1136/amiajnl-2014-002746. PubMed PMID: 24821738; PMCID: PMC4078291.

8. Seid M, Hartley DM, Margolis PA. A science of collaborative learning health systems. Learn Health Syst. 2021;5(3):e10278. Epub 20210601. doi: 10.1002/lrh2.10278. PubMed PMID: 34277944; PMCID: PMC8278442.

9. Finster M, Wenzel M, Taghizadeh E. Common data models and data standards for tabular health data: a systematic review. BMC Med Inform Decis Mak. 2025;25(1):422. Epub 20251113. doi: 10.1186/s12911-025-03267-2. PubMed PMID: 41233809; PMCID: PMC12616946.

10. Wilkinson MD, Dumontier M, Aalbersberg IJ, Appleton G, Axton M, Baak A, Blomberg N, Boiten JW, da Silva Santos LB, Bourne PE, Bouwman J, Brookes AJ, Clark T, Crosas M, Dillo I, Dumon O, Edmunds S, Evelo CT, Finkers R, Gonzalez-Beltran A, Gray AJ, Groth P, Goble C, Grethe JS, Heringa J, t Hoen PA, Hooft R, Kuhn T, Kok R, Kok J, Lusher SJ, Martone ME, Mons A, Packer AL, Persson B, Rocca-Serra P, Roos M, van Schaik R, Sansone SA, Schultes E, Sengstag T, Slater T, Strawn G, Swertz MA, Thompson M, van der Lei J, van Mulligen E, Velterop J, Waagmeester A, Wittenburg P, Wolstencroft K, Zhao J, Mons B. The FAIR Guiding Principles for scientific data management and stewardship. Sci Data. 2016;3:160018. Epub 20160315. doi: 10.1038/sdata.2016.18. PubMed PMID: 26978244; PMCID: PMC4792175.

11. Kahn MG, Callahan TJ, Barnard J, Bauck AE, Brown J, Davidson BN, Estiri H, Goerg C, Holve E, Johnson SG, Liaw ST, Hamilton-Lopez M, Meeker D, Ong TC, Ryan P, Shang N, Weiskopf NG, Weng C, Zozus MN, Schilling L. A Harmonized Data Quality Assessment Terminology and Framework for the Secondary Use of Electronic Health Record Data. EGEMS (Wash DC). 2016;4(1):1244. Epub 20160911. doi: 10.13063/2327-9214.1244. PubMed PMID: 27713905; PMCID: PMC5051581.

12. Marsolo K, Curtis L, Qualls L, Xu J, Zhang Y, Phillips T, Hill CL, Sanders G, Maro JC, Kiernan D, Draper C, Coughlin K, Dutcher SK, Hernandez-Munoz JJ, Falconer M. Assessing the harmonization of structured electronic health record data to reference terminologies and data completeness through data provenance. Learn Health Syst. 2025;9(2):e10468. Epub 20241021. doi: 10.1002/lrh2.10468. PubMed PMID: 40247903; PMCID: PMC12000768.

13. George AP, Powell RA, Rao M. Implementing the English health inequalities agenda: addressing challenges to person-level, cross-sectoral data linkage, access and routine use for local authority public health. Int J Popul Data Sci. 2023;8(4):2166. Epub 20240207. doi: 10.23889/ijpds.v8i4.2166. PubMed PMID: 38481434; PMCID: PMC10933699.

14. Pearce LA, Borschmann R, Young JT, Kinner SA. Advancing cross-sectoral data linkage to understand and address the health impacts of social exclusion: Challenges and potential solutions. Int J Popul Data Sci. 2023;8(1):2116. Epub 20230607. doi: 10.23889/ijpds.v8i1.2116. PubMed PMID: 37670956; PMCID: PMC10476462.

15. Bowker GC, Star SL. Sorting Things Out: Classification and Its Consequences. Cambridge, MA: MIT Press; 2000.

16. Star SL, Ruhleder K, editors. Steps toward an ecology of infrastructure: complex problems in design and access for large-scale collaborative systems. Proceedings of the 1994 ACM Conference on Computer Supported Cooperative Work; 1994.

17. Hernandez AF, Shenkman E, McTigue K, Kepler L, Cohen LW, Jolles MP, Rothman RL, Block JP, Carton TW, Tobin J, Priest EL, Evans C, Heintzman J, Williams DA. PCORnet(R): An Infrastructure Supporting Innovation in Clinical Study Design. Med Care. 2026;64(2S Suppl 3):S178–S84. Epub 20260108. doi: 10.1097/MLR.0000000000002245. PubMed PMID: 41504744; PMCID: PMC12783335.

18. Mandl KD, Kohane IS, McFadden D, Weber GM, Natter M, Mandel J, Schneeweiss S, Weiler S, Klann JG, Bickel J, Adams WG, Ge Y, Zhou X, Perkins J, Marsolo K, Bernstam E, Showalter J, Quarshie A, Ofili E, Hripcsak G, Murphy SN. Scalable Collaborative Infrastructure for a Learning Healthcare System (SCILHS): architecture. J Am Med Inform Assoc. 2014;21(4):615–20. Epub 20140512. doi: 10.1136/amiajnl-2014-002727. PubMed PMID: 24821734; PMCID: PMC4078286.

19. Palinkas LA, Brown CH, Gotham HJ, McGinty EE, Lai E, Adams MCB, McGovern MP. Research Support Networks for Substance Use Translational Research: A Qualitative Evaluation of the HEAL Data to Action (HD2A) Program. Res Sq. 2026. Epub 20260129. doi: 10.21203/rs.3.rs-8340202/v1. PubMed PMID: 41646287; PMCID: PMC12869576.

20. O’Brien BC, Harris IB, Beckman TJ, Reed DA, Cook DA. Standards for reporting qualitative research: a synthesis of recommendations. Acad Med. 2014;89(9):1245–51. doi: 10.1097/ACM.0000000000000388. PubMed PMID: 24979285.

21. Sandelowski M. Whatever happened to qualitative description? Res Nurs Health. 2000;23(4):334–40. doi: 10.1002/1098-240x(200008)23:4<334::aid-nur9>3.0.co;2-g. PubMed PMID: 10940958.

22. Sandelowski M. What’s in a name? Qualitative description revisited. Res Nurs Health. 2010;33(1):77–84. doi: 10.1002/nur.20362. PubMed PMID: 20014004.

23. Hsieh HF, Shannon SE. Three approaches to qualitative content analysis. Qual Health Res. 2005;15(9):1277–88. doi: 10.1177/1049732305276687. PubMed PMID: 16204405.

24. Damschroder LJ, Aron DC, Keith RE, Kirsh SR, Alexander JA, Lowery JC. Fostering implementation of health services research findings into practice: a consolidated framework for advancing implementation science. Implement Sci. 2009;4:50. Epub 20090807. doi: 10.1186/1748-5908-4-50. PubMed PMID: 19664226; PMCID: PMC2736161.

25. Damschroder LJ, Reardon CM, Widerquist MAO, Lowery J. The updated Consolidated Framework for Implementation Research based on user feedback. Implement Sci. 2022;17(1):75. Epub 20221029. doi: 10.1186/s13012-022-01245-0. PubMed PMID: 36309746; PMCID: PMC9617234.

26. Fryer K, Reid CN, Chaphalkar C, Marshall J, Szalacha L, Johnson K, Wright T, Read C, Khan A, Wilson A, Ratani M, Cox K, Tavolieri A, Sampayo M, Su R, Campbell K, Salemi JL. Development of a 5-Step Electronic Medical Record-Based Algorithm to Identify Patients with Opioid Use Disorder in Pregnancy. J Registry Manag. 2024;51(2):69–74. PubMed PMID: 39184206; PMCID: PMC11343436.

27. Mandel JC, Kreda DA, Mandl KD, Kohane IS, Ramoni RB. SMART on FHIR: a standards-based, interoperable apps platform for electronic health records. J Am Med Inform Assoc. 2016;23(5):899–908. Epub 20160217. doi: 10.1093/jamia/ocv189. PubMed PMID: 26911829; PMCID: PMC4997036.

28. Ratta AK, Elger RS, Vermeer W, Wright T, Oxner A, Fryer K, Marshall J. Referral linkages to support pregnant and postpartum individuals with opioid use disorders in Florida: a social network analysis. Addict Sci Clin Pract. 2026;21(1). Epub 20260508. doi: 10.1186/s13722-026-00675-1. PubMed PMID: 42104470; PMCID: PMC13156855.

29. Aarons GA, Hurlburt M, Horwitz SM. Advancing a conceptual model of evidence-based practice implementation in public service sectors. Adm Policy Ment Health. 2011;38(1):4–23. doi: 10.1007/s10488-010-0327-7. PubMed PMID: 21197565; PMCID: PMC3025110.

30. Feldstein AC, Glasgow RE. A practical, robust implementation and sustainability model (PRISM) for integrating research findings into practice. Jt Comm J Qual Patient Saf. 2008;34(4):228–43. doi: 10.1016/s1553-7250(08)34030-6. PubMed PMID: 18468362.

31. Glasgow RE, Vogt TM, Boles SM. Evaluating the public health impact of health promotion interventions: the RE-AIM framework. Am J Public Health. 1999;89(9):1322–7. doi: 10.2105/ajph.89.9.1322. PubMed PMID: 10474547; PMCID: PMC1508772.

32. Woodward EN, Matthieu MM, Uchendu US, Rogal S, Kirchner JE. The health equity implementation framework: proposal and preliminary study of hepatitis C virus treatment. Implement Sci. 2019;14(1):26. Epub 20190312. doi: 10.1186/s13012-019-0861-y. PubMed PMID: 30866982; PMCID: PMC6417278.

